# Impact of the Covid-19 pandemic on elective and trauma orthopaedic surgery in a tertiary referral hospital in Kenya: A retrospective cross-sectional study

**DOI:** 10.1101/2024.12.10.24318827

**Authors:** Njalalle Baraza, Ian Odari, Emmanuel Marsuk Lomole, Sarah Karanja, Mordicai Ating’a

**Affiliations:** Aga Khan University Hospital, Nairobi, Kenya, Department of Surgery; Aga Khan University Hospital, Nairobi, Kenya, Department of Anaesthesia; Kenya Medical Research Institute (KEMRI)

**Author notes:** **Corresponding author:** Njalalle Baraza, FRCS (Tr&Orth).

## Abstract

The impact of the COVID-19 pandemic is still being felt in multiple spheres of life. In healthcare, COVID-19 noticeably affected surgical practice in sub-Saharan Africa indicated by a reduction in elective cases with prioritisation of cancer and emergency cases. This study sought to describe the impact of COVID-19 on orthopaedic surgery volumes in a private tertiary referral hospital.

Ethical approval was obtained for this retrospective cross-sectional study carried out at the Aga Khan University Hospital, Nairobi. The orthopaedic data was collected using the Electronic Healthcare Record (EHR) system in the hospital. Elective and trauma caseloads recorded during COVID-19 were compared with similar quarters in the pre and post COVID-19 periods.

In the third quarter before the COVID-19 pandemic, there were 15 arthroplasty, 53 arthroscopy and 31 implant removal cases. During the pandemic, there was an expected fall in the numbers of procedures by 33%, 30% and 13% respectively compared to the previous (pre pandemic) year. There was also the expected drop in the number of adult trauma cases, however, there was an increase in paediatric fracture fixation with k-wires. In spinal surgery, there was a 71% increase in the number of laminectomies and anterior cervical decompression and fusions (ACDF) from the pre pandemic 21 cases recorded.

The COVID-19 pandemic had an impact on elective and emergency orthopaedic procedure volumes. There was a reduction in the number of elective arthroplasty, arthroscopy, nailing and orthopaedic implant removal procedures. On the other hand there was a significant increase in the number of paediatric fracture fixation and with K-wires and ACDF/laminectomy volumes.

## Introduction

The COVID-19 pandemic, declared a global health crisis by the World Health Organization (WHO) in March 2020, affected healthcare systems throughout the world. Hospitals were forced to reallocate resources, reduce elective procedures, and prioritise the management of COVID-19 cases, which had a profound impact on the availability of surgical services, including orthopaedic care (Al-Jabir et al., 2020). Elective surgeries, in particular, were delayed or cancelled to preserve hospital capacity, while emergency orthopaedic procedures continued but faced new challenges such as limited operating room availability and altered patient care protocols (Pronk, 2020). As it spread, it was branded first as a public health emergency and then a global pandemic (1). Several years on, its impact continues to be felt, particularly in hospitals. With regards to surgical practise in non-specialised centres around the world, the majority of orthopaedic interventions are elective, accounting for up to two-thirds of procedures. These numbers may be different in specialised trauma centres in LMICs, where, due to urbanisation, the number of trauma-related orthopaedic surgeries predominate (2).

The impact of the pandemic on orthopaedic surgery has varied across healthcare systems, depending on local COVID-19 case loads, healthcare policies, and resource allocation strategies. Studies from high-income countries reported significant reductions in elective orthopaedic volumes, with some estimating reductions of up to 50-70% during peak pandemic periods (Bram 2020, Jain 2021). Conversely, emergency orthopaedic volumes were less uniformly affected, with some studies showing a reduction in trauma cases due to lockdowns and reduced mobility, while others reported a surge in trauma cases post-lockdown as normal activities resumed (Mahmood, 2021, Wong, 2021).

However, there is a paucity of data from low and middle-income countries (LMICs) on the impact of the pandemic on orthopaedic surgery volumes, particularly in tertiary care settings that often serve as referral centres for both elective and emergency cases. Understanding the changes in surgical volumes in these settings is crucial to informing future resource allocation, improving patient care pathways, and ensuring system resilience in the face of ongoing or future public health crises.

Although sub-Saharan Africa was one of the least affected regions in terms of morbidity, mortality and disease, the pandemic caused significant disruption of socio-economic life (3). In 2020, the Surgical Society of Kenya developed guidelines with the aim of reducing contamination, optimising resource allocation and aiding surgical decision making during the pandemic (4).

This article aims to assess the impact of the COVID-19 pandemic on both elective and emergency orthopaedic surgery volumes in a tertiary hospital in Kenya. By comparing pre-pandemic and pandemic-period data, this study seeks to provide insight into the magnitude of surgical disruptions and the broader implications for patient outcomes and healthcare resource management.

## MATERIALS AND METHODS

A retrospective cross-sectional study was carried out at the Aga Khan University Hospital, Nairobi, a tertiary referral hospital servicing a wide region including East and Central Africa. The study reviewed the orthopaedic workload before, during and after the covid-19 pandemic. The analysis focused on the 3rd quarters of the three periods to provide a comparison of the orthopaedic trauma and elective workloads and the changes caused by the pandemic. We used the same quarter in each year to limit bias that may have arisen from seasonal variations in either trauma or elective work. We specifically analysed the periods from the beginning of July to the end of September in the years 2019 (before), 2020 (during) and 2022 (after). The data from 2021 was not analysed as this period was still under some COVID-19 restrictions and was not yet a period of normal activity.

All the patients admitted for selected orthopaedic procedures were included in a non-random sequential manner. Patients having either single or multiple procedures in one visit were counted as a single theatre encounter. However, those having multiple procedures in different visits had each encounter counted as a procedure separately. Each procedure was assessed and graded as either elective or trauma emergency.

The data was collected from the electronic health record (EHR) system which was able to identify all the procedures coded as orthopaedic. To avoid missing data that may have been miscoded the review looked at specific operation codes enabling complete data collection even if the cases were under different specialities for example a patient admitted with a general surgery concern but also an orthopaedic problem. These records were then reviewed to determine whether they were performed as elective or trauma admissions. The procedures were then placed into broad categories to allow comparison of representative case types in the study periods (see fig 1). After being categorised, patient identifiers were erased and thus no access to individual patient records by the authors.

In order to extract data from the electronic health record, specific elective and emergency procedures were selected to gauge the effect of the pandemic on practice. For elective surgery, the most commonly performed procedures were selected with the bulk of elective orthopaedic surgery being either joint replacement or arthroscopy. We included removal of metal work as this was a relatively common procedure that was often performed as a day surgery and is relatively inexpensive. Common spine surgery cases were also included to give a representative spread of orthopaedic cases. The selected procedures were arthroplasty, arthroscopy, implant removal, anterior cervical decompression and fusion (ACDF) and laminectomy. For trauma surgery, the procedures selected were open reduction and internal fixation (ORIF)/intramedullary nailing, k-wiring and manipulation under anaesthetic. Both elective and trauma cases that were reviewed are summarised below.

### Elective surgery key orthopaedic procedures

1. Arthroplasty
2. Arthroscopy
3. Implant removal
4. Anterior cervical decompression and fusion (ACDF) and laminectomy

### Trauma and emergent surgery procedures

1. Open reduction and internal fixation (ORIF)/intramedullary nailing.
2. K-wiring.
3. Manipulation under anaesthetic.

Ethical approval for the study was obtained from the hospital Institutional Scientific Ethics Review Committee (ISERC).

### Data Analysis

Data analysis, tables and graphs were performed using Google sheets.

## RESULTS

### Elective

For the categories chosen for analysis, overall there were 120 cases performed before COVID-19, 110 cases during COVID-19 and 131 cases in the post COVID-19 phase in the third quarters. The number of surgical procedures in arthroplasty, arthroscopy and implant removal, saw an expected fall of 33%, 30% and 13% respectively compared to the previous (pre-covid) year. In spinal surgery however, there was a 71% increase in the number of laminectomies and ACDF.

|  | **2019 (pre-covid)** | **2020 (covid)** | **2022 (post-covid)** |
| --- | --- | --- | --- |
| **Arthroplasty** | 15 | 10 | 14 |
| **Arthroscopy** | 53 | 37 | 64 |
| **ACDF/laminectomy** | 21 | 36 | 21 |
| **Implant removal** | 31 | 27 | 32 |

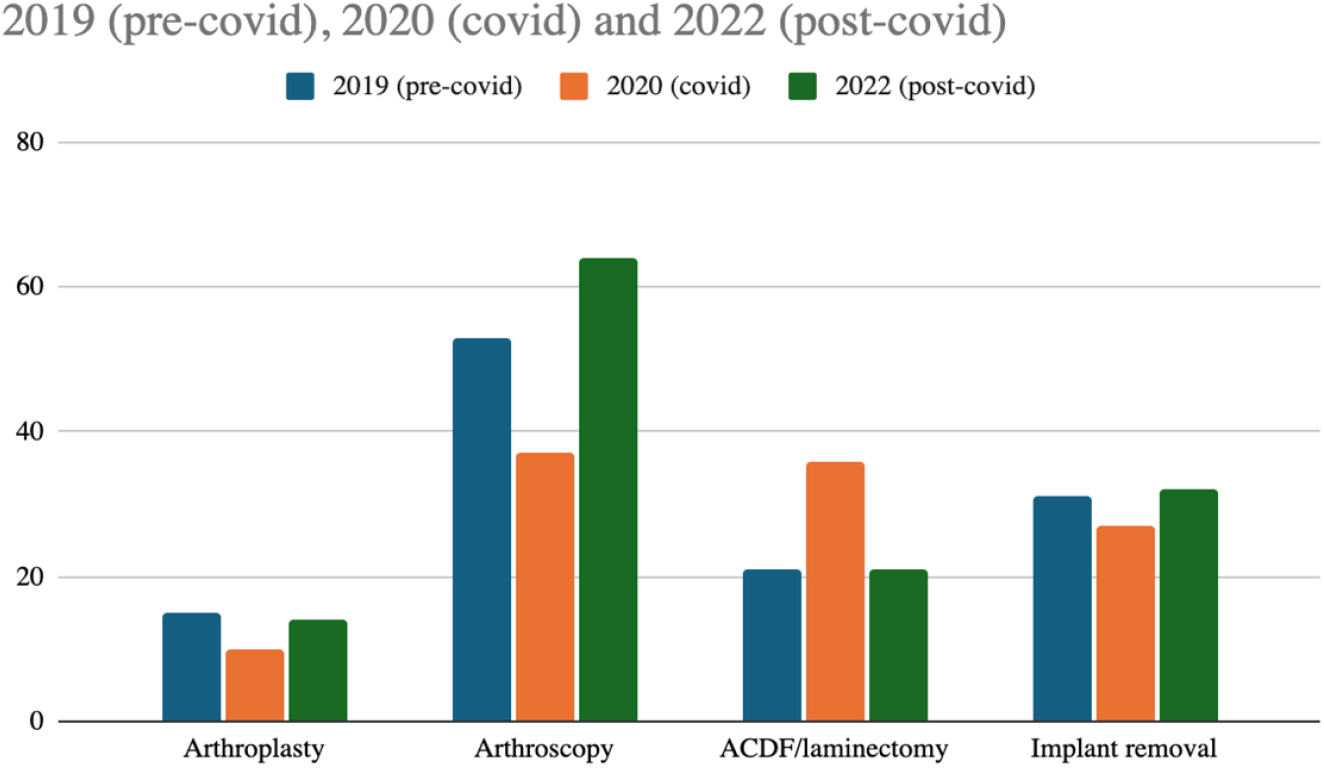

### Trauma

In trauma, there was a reduction in the number of fracture fixations/intramedullary nails and manipulations by 14% and 46% respectively during the covid period. There was, however, a 183% increase in K-wire fixation which, on closer inspection, was all in a paediatric population. This increase slightly reduced after covid-19 pandemic but was still higher than the 2019 baseline by 75%. Fixation/nailing in the post covid period had increased by 26% compared to baseline, while fracture manipulations had reduced by 61% compared to the 2019 baseline.

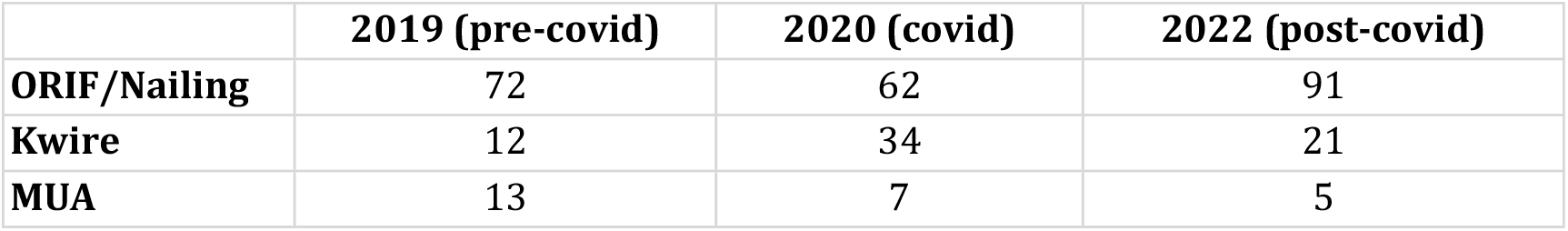

|  | 2019 (pre-covid) | 2020 (covid) | 2022 (post-covid) |
| --- | --- | --- | --- |
| **ORIF/Nailing** | 72 | 62 | 91 |
| **Kwire** | 12 | 34 | 21 |
| **MUA** | 13 | 7 | 5 |

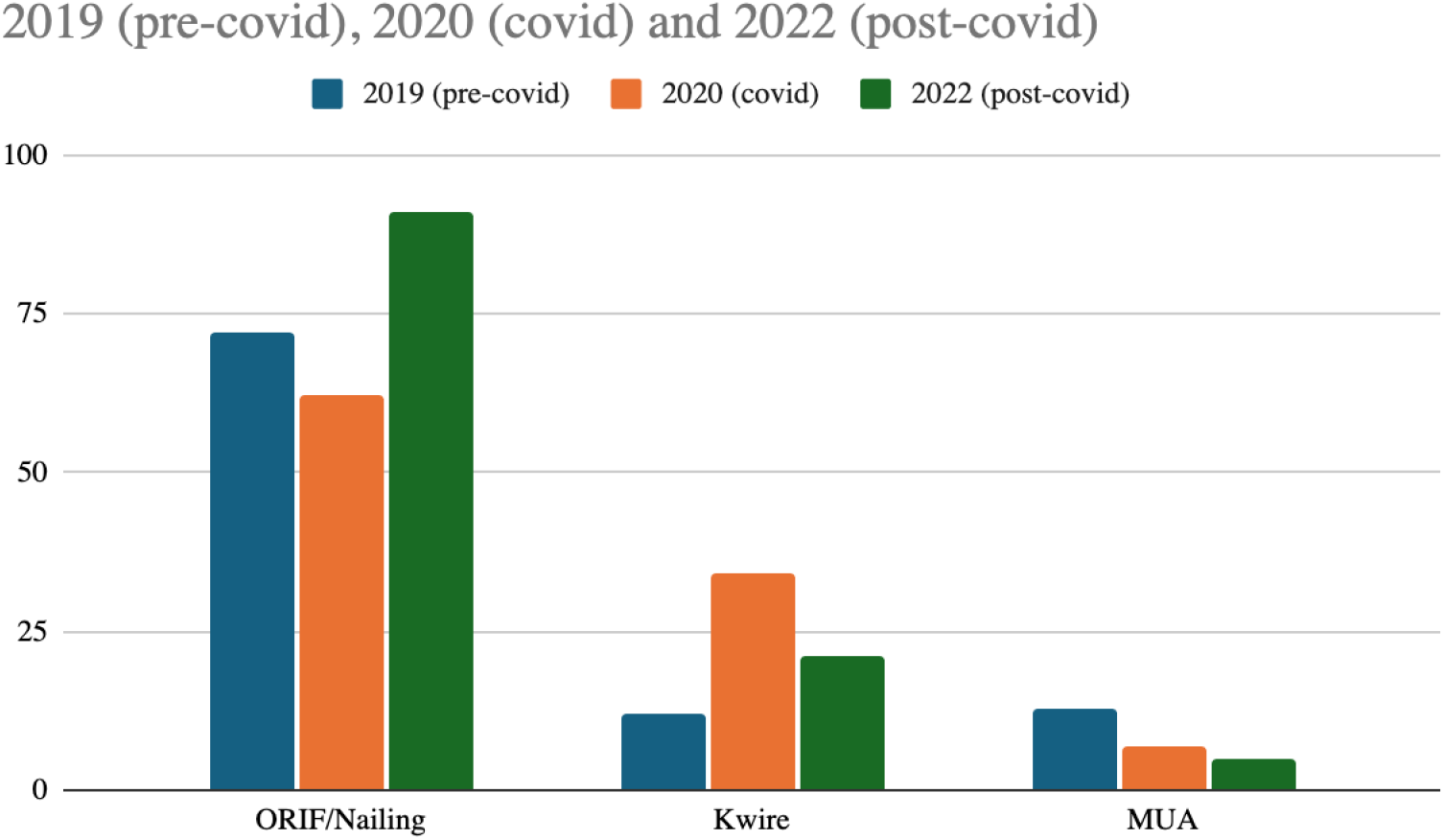

## DISCUSSION

The findings of this study demonstrate that the COVID-19 pandemic had a significant impact on both elective and emergency orthopaedic procedure volumes in our tertiary hospital. Specifically, elective orthopaedic procedures such as arthroplasty, arthroscopy, nailing, and orthopaedic implant removal saw a reduction, while paediatric fracture fixation with K-wires and anterior cervical discectomy and fusion (ACDF)/laminectomy procedures experienced an increase in volumes. These findings are consistent with global trends reported in the literature and highlight the pandemic’s selective impact on different types of orthopaedic procedures. This was in part postulated to be due to increased access to private facilities by holders of the government’s health insurance fund, a measure undertaken by the Kenyan government in response to the crisis, anecdotally to ease the pressure on public sector hospitals. Additionally the spine surgery team pushed for daycare surgery which was attractive to worried patients and reduced costs further. By 2022, the numbers of operations had returned to pre-covid levels with the exception of arthroscopy numbers which had increased, which could be explained by the possible backlog of patients who didn’t have their surgery in the pandemic period or by increased uptake of sporting activities after the pandemic.

The reduction in elective orthopaedic surgeries, including arthroplasty, arthroscopy, and orthopaedic implant removals, aligns with other studies that have documented the widespread deferral of non-urgent surgeries during the pandemic. In many healthcare settings, elective procedures were intentionally delayed to conserve hospital resources, such as ICU beds, ventilators, and personal protective equipment, and to minimise the risk of viral transmission (Pronk, 2021, Al-Jabir, 2020). Arthroplasty procedures, which are typically performed to address chronic degenerative conditions such as osteoarthritis, were particularly affected, as they were classified as non-emergent surgeries during the height of the pandemic (Megaloikonomos, 2020). The decrease in arthroscopy and implant removal procedures can similarly be attributed to their elective nature, where delays would not result in immediate harm to patients (Bram, 2020).

Looking at trauma, there was a reduction in the number of fracture fixations/nailings and manipulations by 14% and 46% respectively during the covid period, which may have been explained by the effect of the curfew that limited night driving, cross country travelling and the restrictions that reduced sports participation. Fixation/nailing in the post covid period had increased by 26% compared to baseline, which could be explained by a resumption of travelling, sports and socialising while manipulations had reduced by 61% compared to the 2019 baseline, perhaps a reflection of perceived end of health austerity measures which had encouraged more conservative practices from the clinicians.

The observed increase in paediatric fracture fixation procedures, particularly using K-wires, reflects a trend seen in some regions where paediatric trauma volumes either remained steady or increased during the pandemic (Jain, 2021,Mahmood, 2021). The likely explanation for this increase lies in the changing patterns of physical activity among children during lockdowns and post-lockdown periods. While organised sports and school activities were restricted, children often engaged in unsupervised play, which may have contributed to an increase in fractures (Wong 2021). In addition, as paediatric fractures are often considered time-sensitive emergencies, the management of these injuries continued with little disruption, in contrast to elective procedures.

One of the most surprising findings of the study was the increase in ACDF/laminectomies and K-wiring during the Covid period compared to the previous year. As was explained earlier, there are various factors which drive healthcare seeking habits, and in our private hospital setting, cost is a major one. Having affordable access to healthcare was the major reason for the spike in ACDF/laminectomy numbers, and is an indirect representation of the effect of cost on health seeking behaviour with patients timing need for intervention with availability of financial resources. These procedures are often performed to address acute spinal conditions, such as cervical myelopathy or disc herniations, that can result in severe pain or neurological compromise if not treated in a timely manner (McCormick, 2020). As a result, these cases were likely prioritized despite the pandemic’s restrictions on elective procedures, as delaying surgery could result in serious functional impairment. This also highlights that even in LMIC, insurance companies and the private sector can play a role in shaping and improving much needed surgical services. Clinician awareness of sensitivity to the general economic climate may have driven the more cost effective K-wires as opposed to other means of fixation (5) and this too represents an interesting area of surgeon influence in the overall cost of healthcare.

We expected that during the pandemic induced lockdown, with reduced travel, sports, socialising and alcohol consumption, there would be a drop in injuries, driving balance of procedures towards elective operating, but the data collected did not reflect this. According to the Kenya national Bureau of statistics data (6), the number of road traffic accidents rose during 2020 with an increase in all categories including, pedestrians, motorcyclists and cyclists. The number of casualties in motorcyclists rose 58% compared to 2019. There was a 17.3% increase in serious injuries involving drivers and an 8% increase in pedestrian casualties. This, however, was not the experience in our tertiary referral hospital where the overall numbers of patients requiring surgery dropped as observed. One possible explanation may have been due to the fact the lockdown restrictions ensured surgical attention was administered in regional facilities with a reduction in transfers to often distant tertiary referral centres.

In a 2021 paper examining the effects of the pandemic on surgical practice, there was found to be a reduction in elective cases and prioritisation of cancer and emergency surgery, and an overall drop in patient volumes, with a rate of cancellations of surgical procedures in the range of 74 to 81% (7). This reduction in elective volumes was in keeping with our findings, as the uncertainty caused by the pandemic may have led patients who had non-urgent conditions to wait until it was deemed safe to return to hospital. Third quarter analysis was used as this was the period that the effect of the pandemic was most felt in the country. With the first case of COVID-19 diagnosed in March 2020 in Kenya, the government put a series of curfews, lockdowns, and movement restrictions in place which had fully taken hold by July 2020. An evaluation of orthopaedic services in other centres around the world showed a reduction as high as 61% in the total number of cases in the second quarter of 2020 compared to the second quarter of the previous year (8). Though their time periods were not precisely the same as ours, there is evidence that although the number of orthopaedics cases performed at our tertiary referral centre followed the general global trend, the consequence was not as profound comparatively.

The emergence of the pandemic highlighted the importance of anticipating such a global crisis. It led to a “mini-revolution” in healthcare with a greater appreciation of the role of technology, i.e., teleconsultation and telerehabilitation. (9). As such, with the trends noted at our unit, we believe that this and similar studies in different settings would be useful in the projection of resource allocation in future similar crises.

Limitations of the study include seasonal variation in clinical workload that was not captured. It would have been valuable to statistically test the difference between the numbers analysed but with no known expected change, comparison groups, or knowledge of numbers who were actually eligible for surgery rather than only those who underwent it, this was not possible. Another potential confounder includes change in surgical personnel over the study period which would have altered the numbers.

Being a cross-sectional retrospective study, confounding factors to the outcomes were not assessed. The study had low external validity as the findings are from a single centre. Ideally, a large multicentre review is needed to fully assess the lasting effects of this pandemic.

## Conclusion

The Covid 19 pandemic had an impact on elective and emergency orthopaedic procedure volumes. There was a reduction in the numbers of common elective procedures including arthroplasty, arthroscopy, nailing and orthopaedic implant removal procedures. On the other hand there was a significant increase in the number of paediatric fracture fixation with K-wires and also in the number of ACDF/laminectomy volumes which reflected school closure and children at home and increased national health insurance funding spending for certain procedures. This study adds to the literature the idea that surgical practice is a distinct resource-intensive health service and we aim to raise awareness of the need to anticipate and innovate in the face of the ever-present possibility of another pandemic.

## Data Availability

Data readily available

